# Myomectomies during cesarean section and outcomes

**DOI:** 10.1101/2024.05.10.24307049

**Authors:** Nikolai N. Ruhliada

## Abstract

The problem of myomectomy during cesarean section is relevant and disputable. Currently, there are no clear guidelines available for conditions and recommendations for performing this procedure during cesarean deliveries. Many obstetricians are hesitant to expand the scope of surgery due to the potential complications, such as increased intraoperative bleeding, postoperative hematomas, the risk of postoperative sepsis, venous thromboembolism, and abdominal adhesions. Additionally, long-term complications can arise from scarring on the uterus that can affect future fertility. This article presents an analysis of case series of cesarean deliveries with and without myomectomies performed at the Department of Obstetrics and Gynecology at St. Petersburg State Medical University in 2022.

## Introduction

Uterine fibroids are a common gynecological disease that affects women of reproductive age. The prevalence of these tumors ranges from 20% to 70% [1,2,3,4]in general gynecologic population and from 0.5% to 6% in pregnant women [5,6,7]. Observed increase in incidence is due to factors such as increasing childbearing age, assisted reproductive technology, hormonal therapy, and contraceptives. Endocrinological disorders also play a role in the occurrence of uterine fibroids. Myoma nodules can grow during pregnancy and are more commonly found in the first trimester [8]. There is currently no consensus on whether a myomectomy should be performed during cesarean section. It is known that the first myomectomy was performed by Victor Bonney, a British gynecologist-surgeon in 1913 in a 30-year-old woman with six large fibroids. After the surgery she gave birth through the natural birth canal three more times [9]. During more than 100 years since this article was published, the situation regarding myomectomies during cesareans has not become clearer. Currently there are no clear guidelines for this procedure, and most Russian and international articles related to the topic present case reports [10,11,12,13,14,15,16]. Opinions about myomectomy during caesarean sections are diverse, some experts believe it can be done safely, while others warn of potential risks, such as increased surgical duration, and complications such as intraoperative bleeding and postoperative sepsis. Long-term concerns include the risk of uterine scarring and its impact on future fertility [17,18,19]. In addition, most obstetricians lack the necessary surgical expertise to perform hysterectomies. The current clinical guidelines from the Ministry of Health in the Russian Federation published in 2020, titled “Uterine Fibroids”, indicate that during caesarian delivery, a myomectomy may be beneficial in case if fibroid prevents the extraction of the fetus [4]. However, practitioners know that uterine contractions after delivery can lead to poor nutrition and ischemia of large fibroids, subinvolution, and impaired emptying of the uterus, which can aggravate the course of the postpartum period. Other recommendations states that myomectomy should be performed during cesareans in cases where certain types of fibroids are present: subserosal nodes located on a thin or broad base, intramural nodules larger than 10 cm with centripetal growth, or if there are multiple fibroids in the cervix or isthmus, or if tumors are malignant; hysterectomy may be recommended [1]. Usually, Krasnopolsky method is used for removing interstitial myoma nodes during a myomectomy [20]. This involves making a linear or oval incision in the uterine tissue with a scalpel until the capsule of the node is visible. Once the capsule is exposed, it is dissected and the tissue of the node pulled into the incision with the forceps. After the node has been removed, the site closed with 2-3 rows of stitches. Disadvantages of this technique include increased bleeding due to uterine changes during pregnancy, difficulties in achieving hemostasis, potential peritoneal damage, and a higher risk of adhesion at the incision site. These factors may lead to longer surgery duration and greater risk for postoperative complications. Literature describes techniques for endometrial myomectomy that can be performed during a caesarean section from within the uterine cavity [21].

The purpose of our study was to evaluate the possibility of performing a myomectomy during cesarean section using various techniques, as well as the effect of increasing the volume of surgery on its duration, intraoperative blood loss and the course of the postoperative period.

## Materials and methods

The study was conducted at the Department of Pregnancy Pathology of the Clinic of St. Petersburg State Pediatric Medical University. We analyzed the protocols of surgeries performed on 377 patients who underwent cesarean sections, as well as the birth records of 69 pregnant women with uterine fibroids. These patients were divided into two groups: Group 1 included 24 patients who underwent myomectomy during cesarean section, while Group 2 included 45 patients who did not have myomectomy (control group).

We reviewed case histories, surgical protocols, and laboratory (general clinical, biochemical, bacteriological, and histological) as well as imaging (ultrasound) data. Statistical analysis of the data was performed using Statistica, and the normality of the feature distribution was determined using the Kolmogorov-Smirnov test with Lilliefors’ modification. Descriptive statistics for normally distributed features are presented as the mean (M) and standard deviation (SD). For the features that do not follow a normal distribution, the median and interquartile range are used. Comparison of sets of quantitative features with a normal distribution is performed using the t-test for independent samples. Features with asymmetric distributions are analyzed using the Mann-Whitney U test. A threshold value of P = 0.05 is used to determine statistical significance. Qualitative data are summarized using absolute and relative frequencies.

## Results

The birth histories of 377 patients of the Department of Pregnancy Pathology of the Perinatal center of the St. Petersburg State Medical University Clinic, who were delivered by caesarean section as planned in 2022, were studied. 69/377 (18.3%) women were diagnosed with uterine fibroids. Myomectomy during cesarean section was performed in 24/377 (6.4%) patients. The decision on the need for myomectomy, taking into account the personalized risks, was made by doctor’s Concilium before the surgery or intraoperatively. Depending on the volume of the operation, the subjects were divided into 2 groups. The first group included 24 patients who underwent myomectomy during cesarean section, the second group consisted of 45 women with uterine fibroids who were delivered by caesarean section on a planned basis without expanding the operation to myomectomy. The average age of patients in the first group was 36.08 (4.4) years versus 38.58 (5.1) years in the second, p = 0.048.

In the group of women who had myomectomy during cesarean section, in 12/24 (50%) uterine fibroids were the main indication for surgical delivery due to the fact that large myoma nodes were located in the isthmus of the uterus along the anterior and posterior walls and would prevent the passage of the fetus through the birth canal, or due to their gigantic size, would potentially lead to labor anomalies. In total, 39 myoma nodes (from 2 to 15 cm) were removed in 24 patients. Most of nodes were located on the anterior wall of the uterine body 23/39 (59.0%), 5/39 (12.8%) nodes were removed from the isthmus of the anterior wall and the fundus of the uterus, 3/39 (7.7%) were located in the isthmus of the posterior wall and in the body of the posterior wall of the uterus. According to the FIGO classification: the vast majority of nodes had an interstitial (type 4) 18/39 (46.1%) and interstitial-subserous (type 5) 14/39 (35.9%) growth pattern. The subserous (type 6-7) type had 5/39 (12.8%) of removed nodes. Nodes with interstitial-submucous (type 2-3) and intraligmental (type 8) growth patterns were less frequent - 1/39 (2.6%) each (Fig 1.).

**Figure 1.**
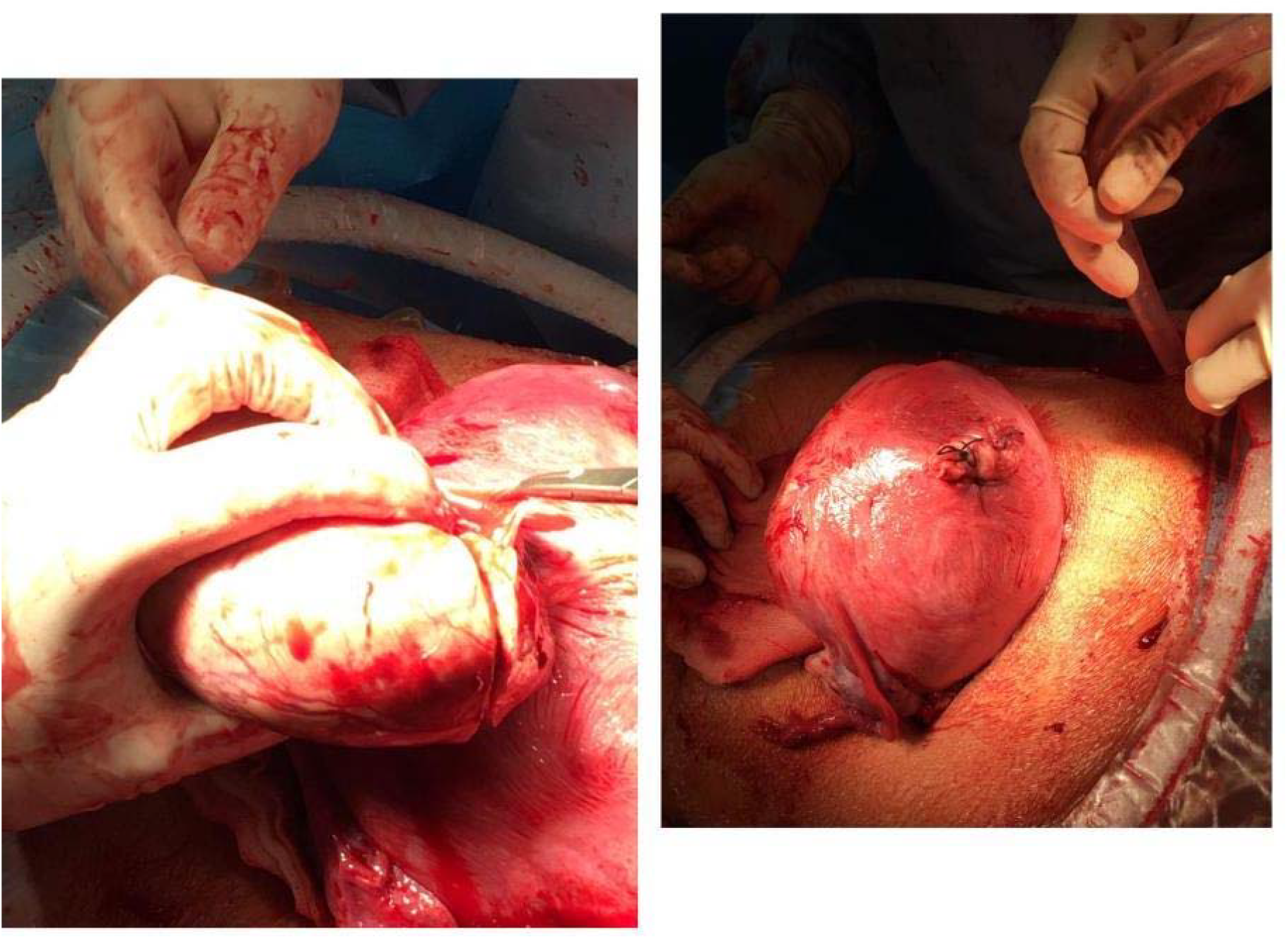
Stages of myomectomy with opening of the serous membrane of the uterus.

The final decision in the surgical technique was made intraoperatively after visual and palpatory assessment of the node. Myomectomy of 2/39 (5.1%) nodes was performed before hysterotomy to extract the fetus. These were cases when large myoma nodes were located in the lower uterine segment of the anterior wall in the area of fetus extraction. However, despite the non-standard start of cesarean section, fetal extraction in these situations occurred at 4 and 5 minutes after the start of the operation and did not affect the newborn’s rate on the Apgar scale at birth. More than half - 22/39 (56.4%) myoma nodes were typically removed after opening the serous membrane of the uterus [20]. However, in these situations, we performed a sagittal incision over the myoma node 2 times smaller than determined node size. The good contractility of the uterus and the reduced size of the incision can reduce the number of possible complications associated with hemostasis in the wound.

Extraction of 2/39 (5.1%) nodes was performed from the side of the uterine cavity (so-called endometrial myomectomy) (Fig. 2). In our opinion, this technique is the most successful for interstitial-submucous and interstitial node growth. The plasticity of the postpartum uterus allows to “squeeze out” the myoma node in a direction most convenient to the surgeon. Operations performed without damaging the uterus serosa look favorably in terms of reducing intraoperative blood loss.

**Figure 2.**
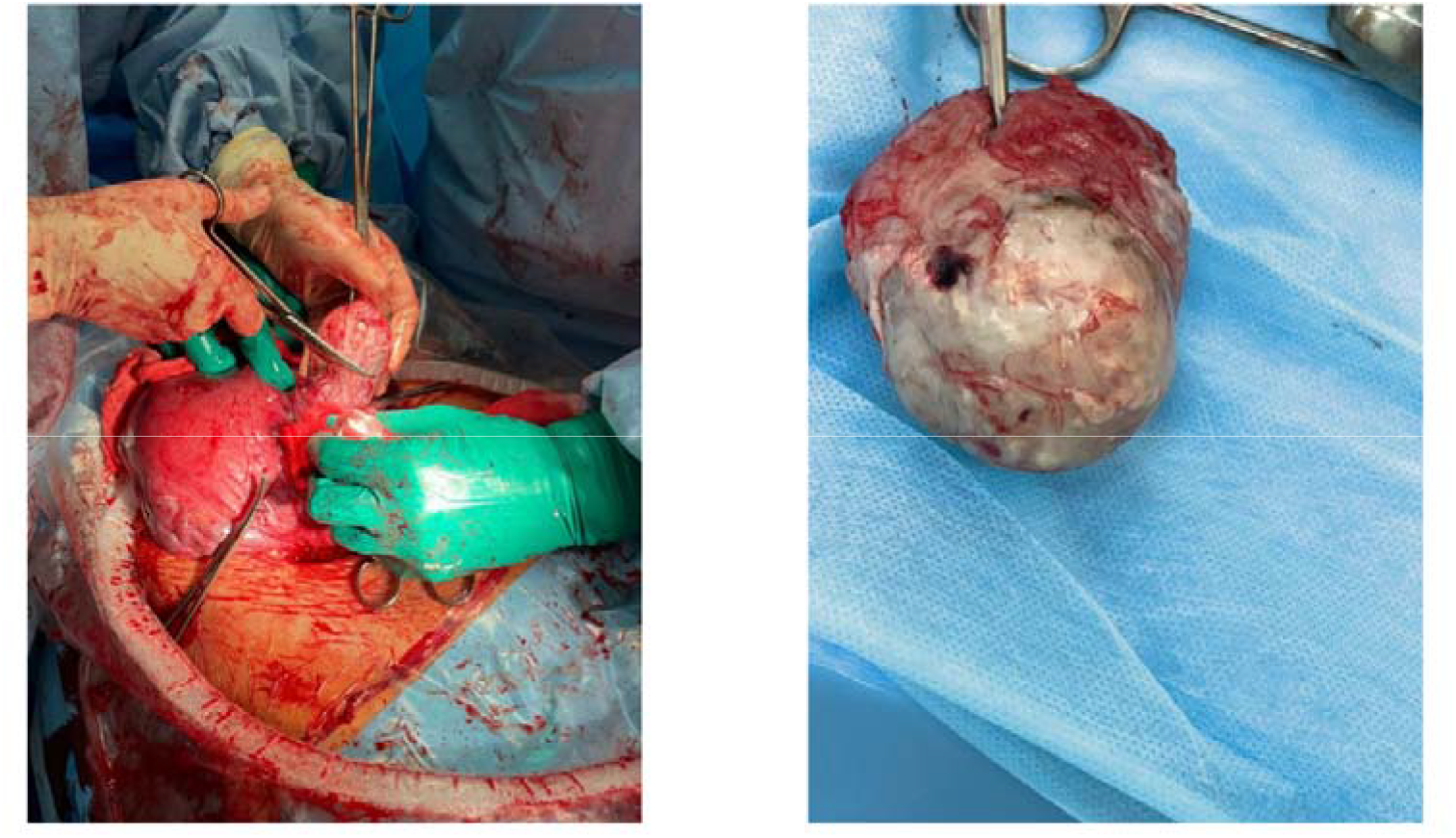
Stages of myomectomy transendometrially (“from inside”).

In 13/39 (33.3%) cases, myomectomy was performed intramurally. This technique was used when the interstitial myoma node was located along the anterior wall of the uterus at a distance of no more than 4 cm from the edge of the incision on the uterus made for cesarean section. Cesarean section is performed typically by a transverse incision in the lower uterine segment. Further, the size, position, and growth pattern of the myoma node are determined visually and by palpation. When a myoma node more than 5 cm size with an interstitial growth pattern (type 3-5 FIGO) is located along the anterior wall of the uterus not more than 4 cm far from the edge of the hysterotomy incision, a transverse incision of the uterine muscle is made in the thickness of the hysterotomy incision made for cesarean section. A tunnel is formed to the lower pole of the myoma node, the node capsule is opened, the node is captured with bullet forceps and enucleated. The length of the incision and the depth of the tunnel depend on the size of the myoma node. The bed of the node and the tunnel made to remove it are sutured through the entire thickness of the uterine wall, with knots tied on the side of the uterus serosa. We believe that performing a myomectomy with intramural access to the myoma node during cesarean section leads to a decrease in uterine traumatization, preservation of the integrity of the uterine serosa, a decrease in the intensity of intraoperative bleeding, reduction of surgery duration, and a decrease in the number of intra- and postoperative complications [22].

In 11/24 (45.8%) women additional surgical procedures were done within cesarean section with myomectomy. In 7/24 (29.2%) patients uterine devascularization by ligation of uterine artery branches was made; 1 case (of 24 (4.2%)) of ovariocystectomy and adhesiolysis each. When performing endometrial myomectomy in one patient, Zhukovsky uterine balloon catheter was used to prevent bleeding in the early postoperative period. A hysterectomy was performed in one patient in myomectomy group. This was in their 30s y.o. patient giving a second birth, with multiple uterine fibroids of large size with an isthmic arrangement of nodes. During the intraoperative revision, it was revealed that the dominant node was 15 cm of size, with an interstitial growth pattern, located in the isthmus of the posterior wall of the uterus. It deformed the uterine cavity due to its size. The second myoma node with a subserous growth pattern and a size of about 8 cm in diameter was located below, in the cervical zone, it had limited mobility. Taking into account the high risk of massive intraoperative bleeding due to the topography and size of the nodes, it was decided to perform a total hysterectomy without uterine appendages (Fig. 3).

**Figure 3.**
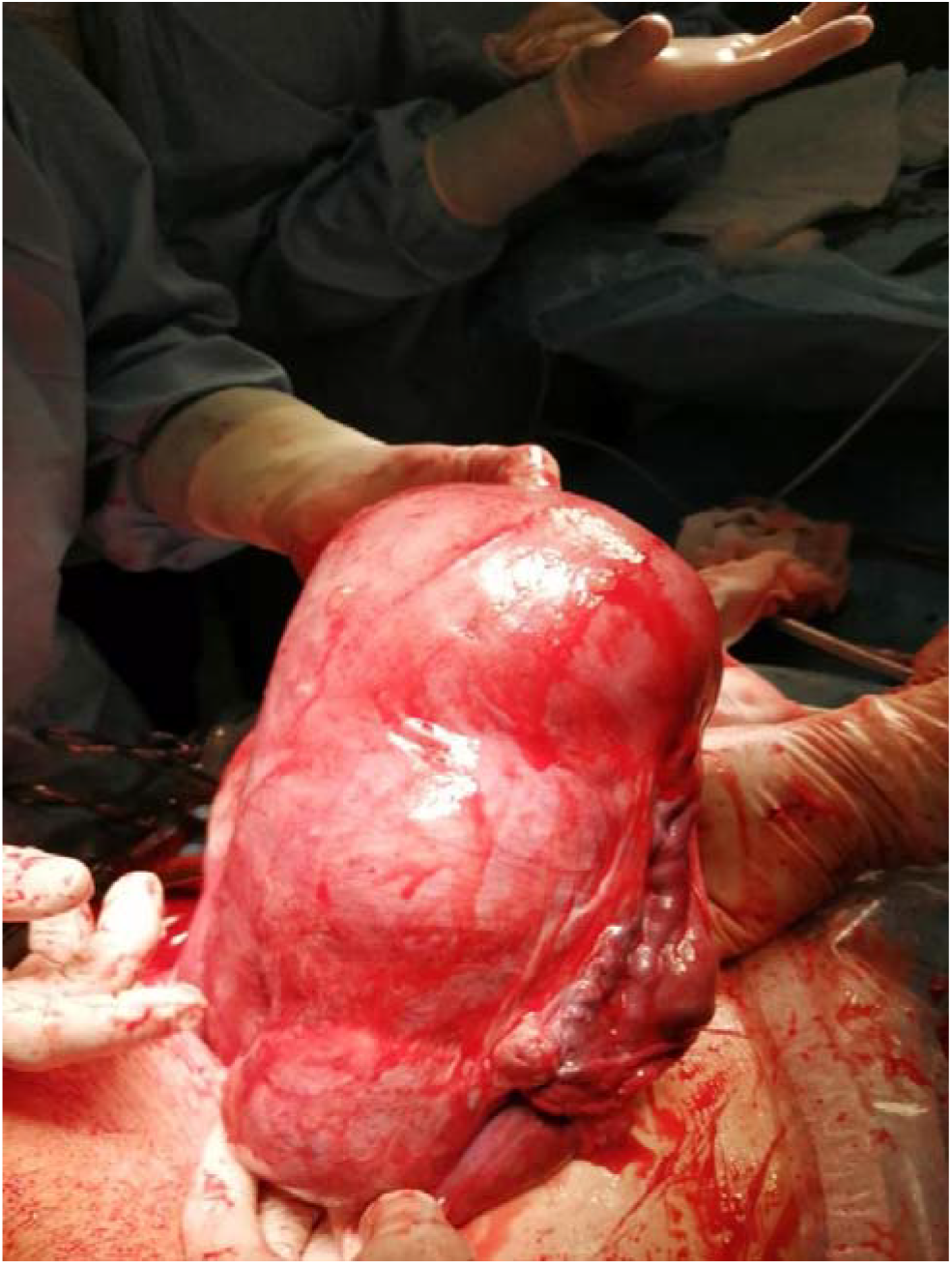
Case for uterus removal.

Within the analysis of operations outcomes, we assessed the duration of surgical interventions, extraction time, blood loss, and the course of the postoperative period in patients of group 1 (with myomectomy during cesarean section n=24) and group 2 (with uterine fibroids and without myomectomy during cesarean section, n=45) (table). It was found that the duration of operation in f the first group was significantly higher: 60.0 minutes (20.49) versus 46.2 (13.15) in the second group (p=0.001) (Table 1). In the second group, 6 pregnant women delivered twins so 51 children were born in 45 patients. There were no statistically significant differences in fetal extraction time from the beginning of the operation; it was 3 (3; 5) minutes in the first group and 4 (3; 5) minutes in the second, (p=0.31). There was no significant difference in calculated volume of intraoperative blood loss: 700 (550; 850) ml in the first group and 650 (500; 700) ml in the second (p=0.64). Uterine fibroids did not have a significant effect on the weight and condition of newborns, children were born without hypoxia with the weight of 3144.17g (452.5) in the first group and 3037.45 g (584.6) in the second (p=0.43). In the postoperative period, all patients from the first group were given antibiotic therapy (ceftriaxone at a daily dose of 2.0 g for 5 days) and venous thrombosis prophylaxis with low-molecular-weight heparins. The volume of the operation did not lead to postoperative complications; the patients of both groups were discharged at 4^th^ -6^th^ day of the postpartum period.

**Table 1.** Comparative characteristics of clinical and anamnestic indicators and characteristics of operations in comparison groups.

| Option | Group 1 (patients with uterine fibroids who underwent cesarean section and myomectomy) | Group 2 (patients with uterine fibroids delivered by cesarean section) | Statistical significance in comparison groups, p |
| --- | --- | --- | --- |
| Age, years M (SD) | 36,08 (4,4), n=24 | 38,58 (5,1) n= 45 | p=0,048 |
| Duration of the operation, min M (SD) | 60,0 (20,49), n=24 | 46,2 (13,15) n= 45 | p=0,001 |
| Fetal extraction time, min Me (Q1;Q3) | 3 (3;5), n=24 | 4 (3;5), n= 51 | p=0,31 |
| Newborn's weight, g, M (SD) | 3144,17 (452,5), n=24 | 3037,45 (584,6), n= 51 | p=0,43 |
| The volume of blood loss, ml Me (Q1;Q3) | 700 (550;850), n=24 | 650 (500; 700), n= 45 | p=0,64 |

Note: To identify statistically significant differences in the normal distribution of features, the analysis was carried out using the T-test of independent samples, data with an asymmetric distribution were analyzed using the Mann-Whitney U-test.

## Conclusion

Myomectomy during abdominal delivery does not lead to increased blood loss or other intraoperative or postoperative complications. The surgery should be performed by a multidisciplinary team which includes surgeons (skilled in hysterectomy, blood vessels ligation, and various types of hemostatic sutures), an anesthesiologist, a transfusion specialist, and a neonatal specialist. Prior to surgery, a careful evaluation including ultrasound and magnetic resonance imaging is necessary to accurately determine the location of the tumor. Patients should be fully informed about the details of the procedure, including the risks and possible complications, and should give consent for a caesarean section with a possible hysterectomy.

Having the risk of intraoperative bleeding, it is important to prepare blood components in advance for potential blood transfusions, use blood-saving techniques such as intraoperative reinfusion, and employ surgical hemostatic measures.

The method of myomectomy is determined by the location of the myoma, its growth pattern, and the surgeon’s preference. Myomectomy may be performed through a surgical incision either intramurally (through the wall of the uterus), typically with the opening of the serous layer above the myoma and subsequent removal of the tumor, or transendometrially.

Our study has confirmed the feasibility and safety of performing myomectomy during a caesarean section, which leads to improved immediate and long-term outcomes and offers significant clinical and economic benefits.

## Data Availability

All data produced in the present study are available upon reasonable request to the authors

